# A Digital Twin for Tracking and Forecasting Glycemia with Septic Patients in ICUs

**DOI:** 10.64898/2026.04.24.26351177

**Authors:** Xiang Cao, Chao Cai, Jianguo Hou, Xinhang Wei, Qi Wang

## Abstract

We present a digital twin framework for real-time glucose monitoring and forecasting in septic patients in intensive care units (ICUs). The framework combines advanced machine learning models trained on continuous glucose measurements with a dynamic transfer-learning workflow that enables rapid deployment to individual patients and supports personalized, adaptive, and predictive clinical decision-making. Built on a foundation model—a pretrained time-series transformer—the digital twin continuously updates its parameters as new patient data arrive and produces rolling near-term forecasts in real time. To assess adaptability and computational efficiency, we deployed the pretrained model to ten septic patients and evaluated multiple retraining strategies, including zero-shot inference, linear probing, and full and staged fine-tuning. Results show that the model can be initialized and personalized for a new patient within seconds on a standard laptop while achieving accurate glucose forecasts under varying data conditions. These findings demonstrate the feasibility of real-time model personalization in resource-constrained, high-acuity environments and highlight the potential of digital twins as scalable, AI-enabled platforms for continuous physiological monitoring, clinical decision support, and individualized treatment design in the ICU.

## 1 Introduction

Sepsis, a life-threatening syndrome caused by a dysregulated host response to infection, remains one of the greatest challenges in intensive care. It is defined by acute organ dysfunction driven by systemic inflammation and microcirculatory failure [18] and continues to be a leading cause of ICU mortality worldwide despite advances in early detection and supportive care [12]. Its clinical presentation is highly heterogeneous, shaped by infection site, host co-morbidities, and treatment timing [1]. This variability, coupled with an incomplete mechanistic understanding, has hindered the development of universal therapies and underscored the need for individualized, adaptive management strategies [5].

Glucose dysregulation is a hallmark of sepsis. Stress-induced elevations of epinephrine, cortisol, and glucagon drive glycogenolysis and gluconeogenesis, producing hyperglycemia, insulin resistance, and oxidative stress that amplify inflammation [11, 14, 15]. Both hyperglycemia and hypoglycemia are associated with increased mortality and delayed recovery [17, 20, 22], yet the optimal therapeutic range remains uncertain [8,19]. Tight glucose control reduces mortality but risks hypoglycemia; moderate control may fail to prevent inflammatory complications. These trade-offs highlight the urgent need for personalized, data-driven glucose management in critical care.

Advances in artificial intelligence (AI) have enabled predictive modeling from continuous monitoring and electronic health records [6, 21]. In healthcare, the emerging digital twin (DT) paradigm—an in-silico replica that synchronizes with real-time patient data—integrates mechanistic understanding with AI-based inference to forecast patient-specific trajectories and guide clinical decisions [7, 13]. In ICUs, where patient states evolve rapidly, a glucose-focused digital twin could anticipate metabolic fluctuations, warn of impending instability, and inform insulin and nutrition adjustments through closed-loop feedback.

Data-driven models now complement classical mechanistic approaches such as the Bergman minimal model [2, 3, 10], overcoming their limitations in generalizability and parameter sensitivity. Deep learning architectures—including continuous-time recurrent neural networks [9] and transformer-based models such as PatchTST [16]—capture nonlinear and long-range dependencies in physiological data, enabling real-time, individualized forecasting.

In this paper, we introduce a digital twin framework for continuous glucose monitoring and forecasting in septic patients in intensive care units (ICUs). Built on a pretrained PatchTST model, the framework incorporates a transfer-learning workflow for rapid patient-specific adaptation and dynamic real-time updating. Leveraging high-resolution continuous glucose monitoring data, we benchmark multiple adaptation strategies to evaluate predictive accuracy, computational efficiency, and practical clinical usability. The resulting digital twin can be individualized within seconds while achieving strong forecasting performance on readily available hardware such as a standard laptop.

Beyond its immediate application, this work provides a concrete development paradigm for digital twins targeting acute medical conditions through a single clinically meaningful physiological variable. By converting continuous data streams into actionable predictive information and coupling forecasting with adaptive learning, the framework moves AI-enabled precision critical care closer to real-time implementation. It further lays the groundwork for scalable multimodal digital twins capable of integrated multi-organ monitoring, decision support, and treatment optimization in the ICU.

## 2 Data Acquisition

### 2.1 Dataset for building the foundation model

The data for building the foundation model (or the template) are obtained from a woman in her 30s with sepsis and diabetes, who presented with sudden dysarthria, convulsions, and loss of consciousness and was admitted to Sichuan Provincial People’s Hospital [4]. Following enrollment, she is fitted with a continuous glucose monitoring (CGM) device (FreeStyle Libre, Abbott Diabetes Care Ltd., UK). Glucose data from the preceding 24 hours are downloaded each morning at 8:00 AM. Upon discharge from the emergency intensive care unit (EICU), the device is removed and stored for centralized data collection. The dataset comprises minute-by-minute glucose measurements collected in December of 2023 for two weeks, yielding 19,621 data points.

### 2.2 Dataset of ten additional septic patients

The overall clinical data for the 10 new patients used in this study include demographics, admission characteristics, and clinical scores (APACHE II, SOFA, and NRS-2002) obtained from the Emergency Medical Record System. Laboratory and biochemical data are retrieved from the Laboratory Information System, and nutritional and insulin administration records are obtained from the Hospital Information System. Continuous physiological variables, including ventilation, infusion, and ECG data, are captured in real time via the EICU automated monitoring platform. The CGM system provides high-resolution, minute-by-minute glucose readings throughout the ICU stay, downloaded daily and archived upon discharge. As shown in [4] that the insulin data has limited impact on the forecasting of the glucose level, we develop digital twins for the ten patients using the CGM-collected glucose data only in this study.

The data collection and its use in this study is part of an observational study, and the study protocol was approved by the Medical Ethics Committee of Sichuan Provincial People’s Hospital and Sichuan Academy of Medical Sciences (approval number: LunShen(Yan)-2026-250) in March, 2026. Written informed consent was obtained from patients or their representatives if the patient was unconscious. The study protocol was registered prior to the study’s commencement at the Chinese Clinical Trial Registry (ChiCTR), registration number: ChiCTR2300077594. The entire study procedure adheres to the principles of the Declaration of Helsinki for medical research involving human participants.

## 3 Methods

### 3.1 Statistical analysis of the datasets

The first aim of the study is to transfer the foundation model, pretrained on the glucose dataset from a previous study [4] using a CGM-collected dataset from a septic patient (called the source dataset), to a new septic patient. The source dataset comprises 19,621 glucose readings from a bedside CGM on the specific septic patient, recorded minute by minute in a two week’s span in December of 2023. It was partitioned into training (70%), validation (10%), and testing (20%) sets for machine learning. The data and the 7:1:2 partition is visualized in Figure 3.1. Subsequently, within each of these partitions, we employ a sliding window technique to generate input-target samples for training the model.

**Figure 3.1:**
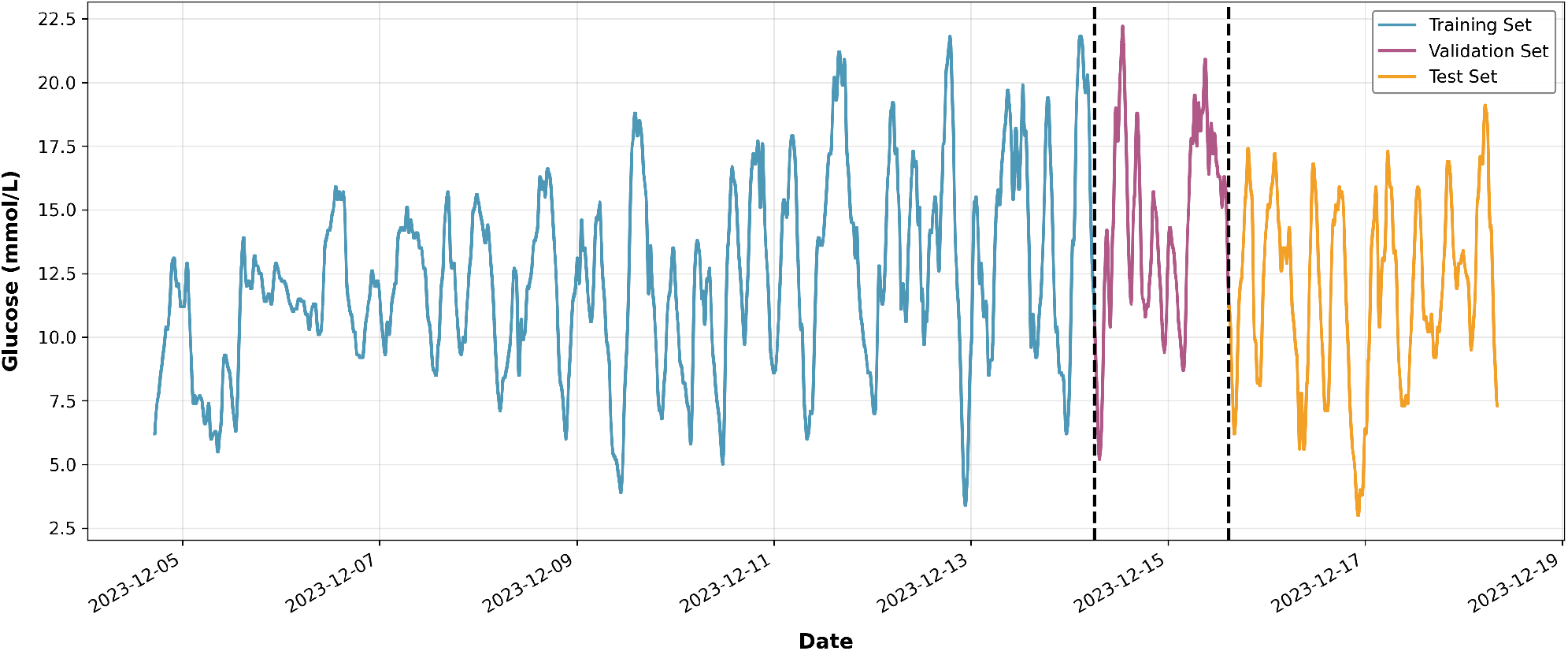
The time series of glucose in the source dataset marked with 7:1:2 data divisions. The dataset is divided into the training (blue, 70%), validation (purple, 10%), and testing (orange, 20%) set. The foundation model is developed using the training and validation data, with final performance evaluated on the testing set.

Specifically, a window of 45 consecutive minutes is used to create a sample. The first 30 minutes in this window serves as the input sequence (look-back window), and the next 15 minutes serves as the target sequence (prediction horizon). The window is moved one minute forward across the time series to generate the next sample. This procedure results in 13,675 training samples, 1,934 validation samples, and 3,895 test samples from the source dataset (see [4]). The predictive model trained on the training and validation sets had been tested on the testing set in [4] and shown to produce excellent predictions in the prediction horizon. It then serves as the template for deploying to a new patient through a newly developed transfer learning workflow in this study. It is therefore termed the foundation model of the digital twin in this paper.

In this investigation, we consider deploying the foundation model to 10 new septic patients via several transfer learning strategies. The number of samples in each category for each new patient will vary from patient to patient however. A statistical summary of these patients, contrasted with the source data, is presented in Table 3.1. A visualization of the time series for the first five of these patients is provided in Figure 3.2. Notably, datasets of new patients are considerably shorter and, in some cases, exhibit distinct data distributions compared to the source dataset for the development of the foundation model. Following the same machine-learning procedure, each patient’s glucose dataset is split into the training, validation, and testing set in a 7:1:2 ratio, respectively. In all the deploying/refitting processes, we configure the model with a 30-minute look-back window and a 15-minute prediction horizon, a setting that has shown to be optimal for glucose forecasting in our previous work for individual patients [4]. The predictive results with the prediction horizon being extended to extended, 30 minutes will be studied in subsequent sections.

**Figure 3.2:**
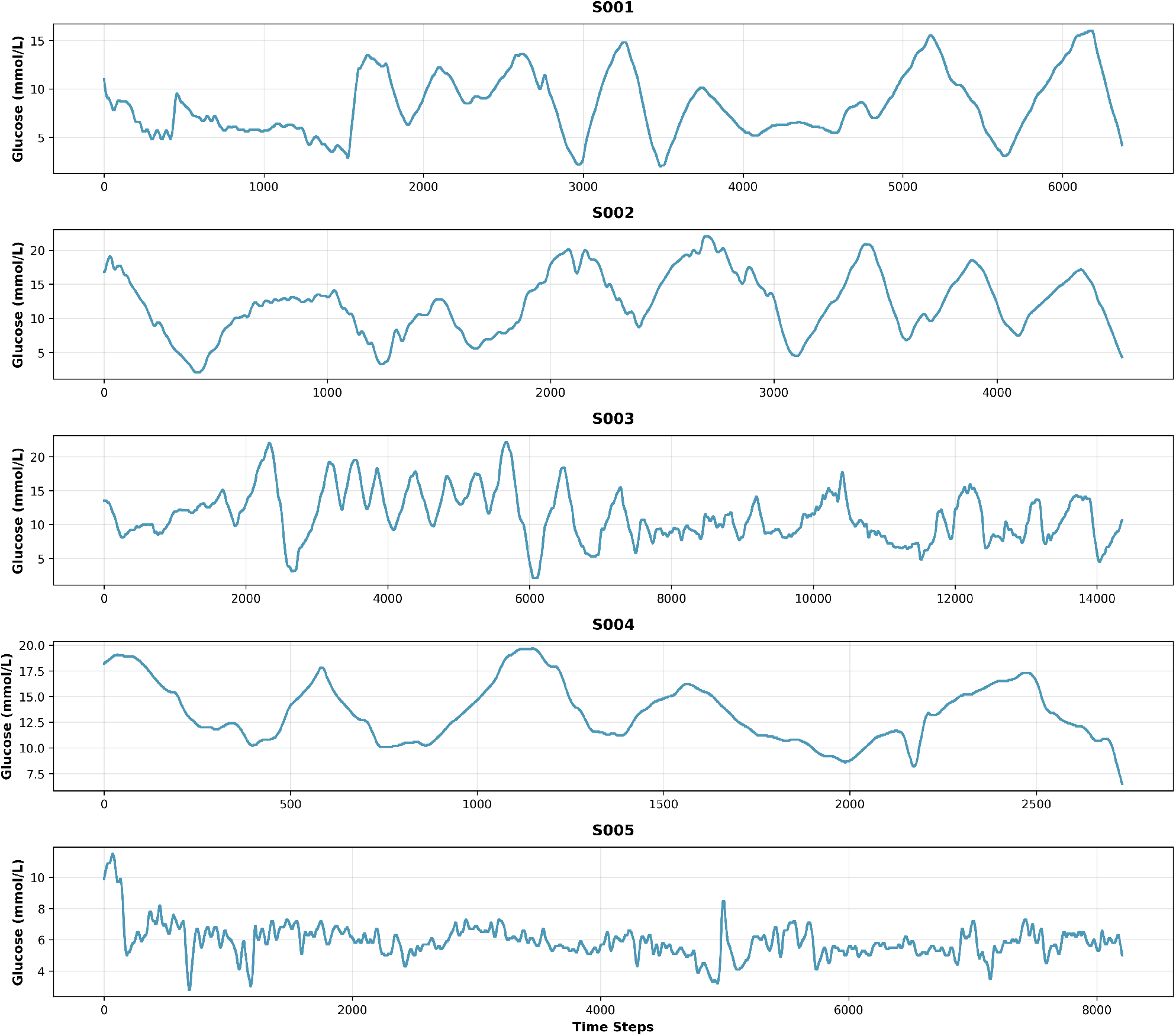
Time series of glucose levels of five randomly selected patients (S001–S005).

**Table 3.1:** Summary statistics of the source and other patients’ glucose datasets.

| Patient | Length | Mean | Std. Dev | Min | Q1 (25<sup>th</sup>) | Q2 (50<sup>th</sup>) | Q3 (75<sup>th</sup>) | Max |
| --- | --- | --- | --- | --- | --- | --- | --- | --- |
| Source | 19,621 | 12.4 | 3.7 | 3.0 | 9.9 | 12.2 | 15.0 | 22.2 |
| S001 | 6,375 | 8.3 | 3.2 | 2.0 | 5.9 | 7.8 | 10.6 | 16.0 |
| S002 | 4,561 | 12.2 | 4.6 | 2.1 | 8.8 | 12.4 | 15.7 | 22.0 |
| S003 | 14,356 | 11.3 | 3.7 | 2.1 | 8.7 | 10.7 | 13.7 | 22.1 |
| S004 | 2,731 | 13.5 | 2.9 | 6.5 | 11.1 | 13.0 | 15.6 | 19.7 |
| S005 | 8,206 | 5.9 | 1.0 | 2.8 | 5.3 | 5.8 | 6.4 | 11.5 |
| S006 | 6,706 | 14.0 | 3.1 | 7.4 | 11.6 | 13.9 | 16.1 | 22.0 |
| S007 | 3,436 | 13.1 | 2.4 | 6.2 | 11.4 | 13.7 | 15.0 | 16.6 |
| S008 | 826 | 13.4 | 3.4 | 7.0 | 11.7 | 12.5 | 14.4 | 21.3 |
| S009 | 1,081 | 11.0 | 1.5 | 7.2 | 10.1 | 11.5 | 12.0 | 14.5 |
| S010 | 1,726 | 14.3 | 2.9 | 8.4 | 12.1 | 14.5 | 16.9 | 19.4 |

### 3.2 Models

Guided by the comprehensive study in [4] for the “best available” architecture of the foundation model, we adopt the Patch-based Time Series Transformer (PatchTST) model, originally developed by Nie et al. [16]. The innovation of PatchTST lies in its ability to efficiently segment time series data into patches to enhance the long-term forecasting accuracy. This model was shown to be the best-performing model on the source dataset for forecasting in [4].

We utilize the PatchTST model pretrained as described in [4] as the foundation model. All experiments for new patients were configured with a 30-step look-back window and a 15-step prediction horizon. To adapt the foundation model to datasets of the new patients, we compare the following five training and fine-tuning strategies.

1. Zero-shot inference: The pretrained model is deployed directly to the new patient dataset without any fine-tuning.
2. Full fine-tuning: All model parameters are updated by retraining on the new dataset with the parameters of the pretrained model as the initial conditions.
3. Linear probing: The model’s backbone is frozen, and only the final output layer is retrained.
4. Staged fine-tuning: It is a two-stage approach where we first train only the output layer for 10 epochs and then fine-tune all parameters for another 10 epochs.
5. Training from scratch: An identical architecture of the neural network model as that of the foundation model’s is trained from a random initialization.

All strategies, except for the staged fine-tuning approach, are trained for 10 epochs. To benchmark their performance, we employ six commonly used metrics: three for absolute errors and three for relative errors, defined in the following.

MSE (Mean Squared Error):

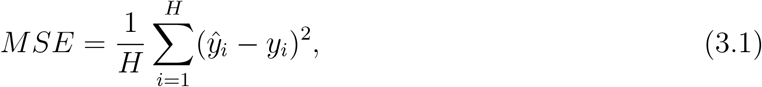

where ŷ_*i*_ is the predicted value, *y*_*i*_ is the value of the ground truth, and *H* is the length of the predicted time horizon.

MAE (Mean Absolute Error),

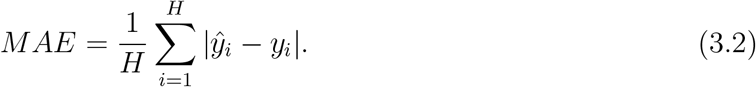

MME (Mean Maximum Error),

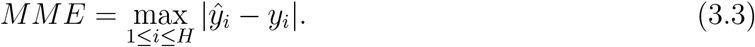

MSPE (Mean Squared Percentage Error),

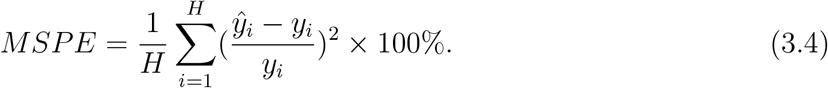

MAPE (Mean Absolute Percentage Error),

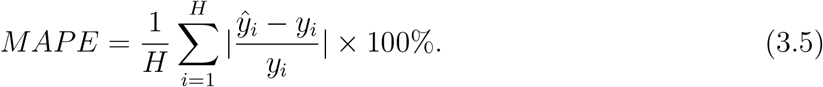

and MMPE (Mean Maximum Percentage Error),

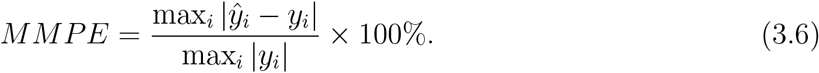

The results for each retraining strategy, including the performance metrics and the corresponding training time, are summarized in Tables 4.1 and 4.2, respectively.

**Table 4.1:** Forecasting performance and computational cost of five training and fine-tuning strategies for patients S001–S005. All experiments are conducted using a 30-step look-back window and a 15-step prediction horizon. The best result is highlighted in bold in each metric for each patient.

| Patient | Method | MSE | MAE | MME | MSPE (%) | MAPE (%) | MMPE (%) | Time (s) |
| --- | --- | --- | --- | --- | --- | --- | --- | --- |
| S001 | Zero-shot | 0.012 | **0.079** | 0.167 | 0.025 | 0.958 | 1.969 | 0.0 |
|  | Finetune | **0.012** | 0.079 | **0.167** | **0.023** | **0.948** | **1.933** | 13.9 |
|  | Linear probe | 0.013 | 0.082 | 0.172 | 0.025 | 0.994 | 2.008 | 8.4 |
|  | Two-stage | 0.013 | 0.081 | 0.170 | 0.023 | 0.966 | 1.968 | 21.3 |
|  | Scratch | 0.016 | 0.088 | 0.183 | 0.025 | 1.040 | 2.075 | 13.9 |
| S002 | Zero-shot | **0.024** | **0.110** | **0.234** | **0.018** | **0.921** | **1.883** | 0.0 |
|  | Finetune | 0.031 | 0.123 | 0.266 | 0.025 | 1.056 | 2.162 | 12.5 |
|  | Linear probe | 0.031 | 0.124 | 0.268 | 0.025 | 1.054 | 2.177 | 6.8 |
|  | Two-stage | 0.031 | 0.121 | 0.267 | 0.026 | 1.042 | 2.178 | 18.4 |
|  | Scratch | 0.027 | 0.113 | 0.253 | 0.023 | 0.980 | 2.105 | 12.1 |
| S003 | Zero-shot | 0.023 | 0.100 | 0.218 | 0.028 | 1.083 | 2.301 | 0.0 |
|  | Finetune | 0.021 | **0.097** | 0.210 | 0.027 | **1.052** | 2.208 | 31.5 |
|  | Linear probe | 0.022 | 0.099 | 0.215 | 0.027 | 1.069 | 2.262 | 17.6 |
|  | Two-stage | **0.021** | 0.097 | **0.210** | **0.027** | 1.055 | **2.205** | 47.9 |
|  | Scratch | 0.025 | 0.107 | 0.222 | 0.030 | 1.136 | 2.297 | 30.9 |
| S004 | Zero-shot | 0.032 | 0.117 | 0.251 | 0.023 | 0.907 | 1.877 | 0.0 |
|  | Finetune | 0.030 | 0.114 | 0.238 | 0.021 | 0.876 | 1.767 | 6.5 |
|  | Linear probe | 0.030 | 0.112 | 0.240 | 0.022 | 0.876 | 1.802 | 4.0 |
|  | Two-stage | **0.028** | **0.112** | **0.232** | **0.020** | **0.858** | **1.723** | 10.0 |
|  | Scratch | 0.064 | 0.149 | 0.309 | 0.047 | 1.178 | 2.341 | 6.7 |
| S005 | Zero-shot | 0.027 | 0.109 | 0.237 | 0.087 | 1.929 | 4.063 | 0.0 |
|  | Finetune | **0.021** | 0.100 | 0.211 | **0.068** | 1.772 | 3.614 | 17.8 |
|  | Linear probe | 0.022 | 0.102 | 0.215 | 0.072 | 1.803 | 3.692 | 10.6 |
|  | Two-stage | 0.021 | **0.099** | **0.210** | 0.069 | **1.765** | **3.600** | 28.1 |
|  | Scratch | 0.026 | 0.110 | 0.229 | 0.083 | 1.944 | 3.922 | 17.8 |

**Table 4.2:**
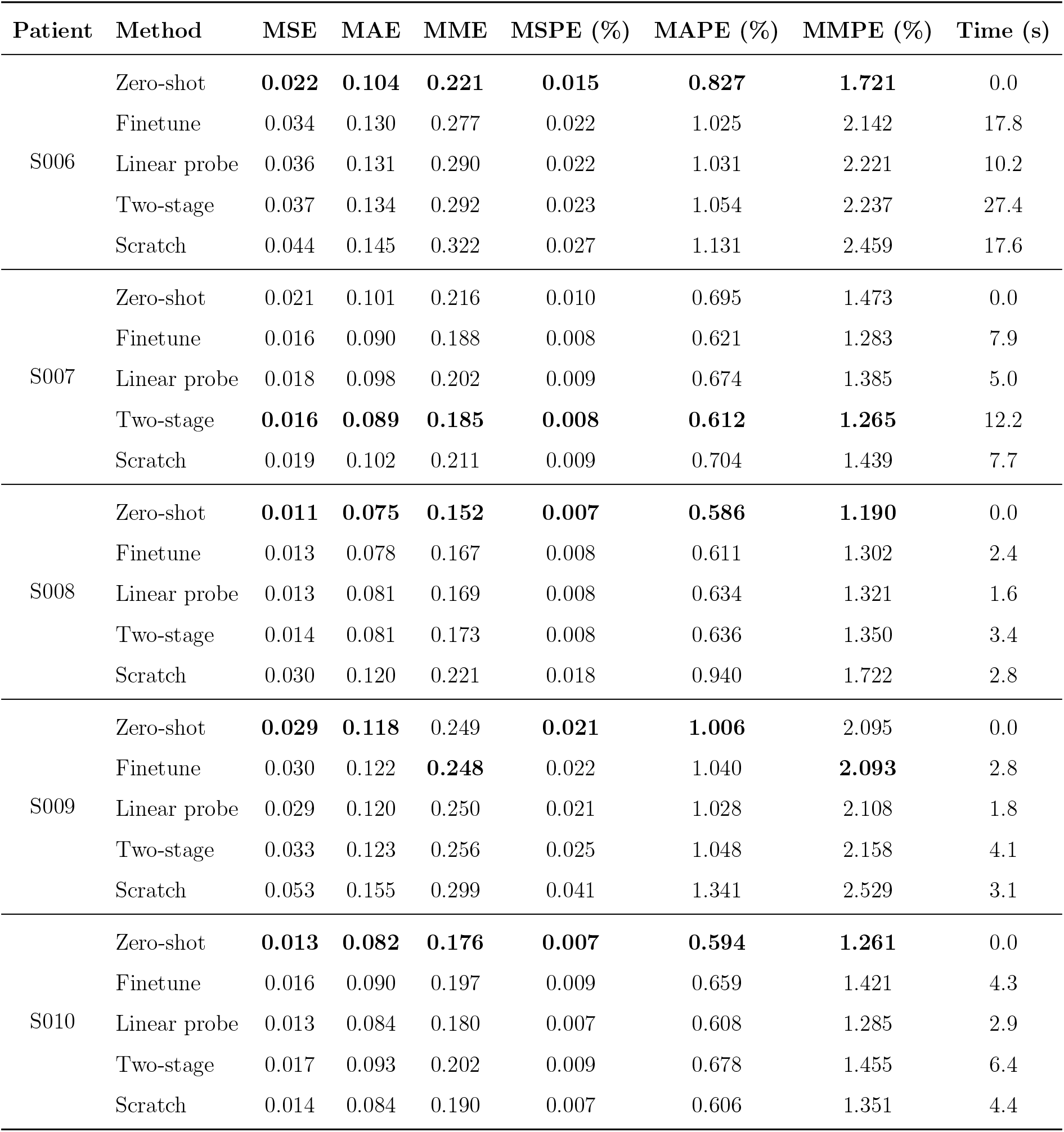
Forecasting performance and computational cost of five training and fine-tuning strategies for patients S006–S010. All experiments are conducted using a 30-step look-back window and a 15-step prediction horizon. The best result is highlighted in bold.

## 4 Results

### 4.1 Evaluation of the retraining strategies

We evaluated five training and fine-tuning strategies for the forecasting task with a look-back window of 30 and a prediction horizon of 15 time steps/units (mins). Except for the zero-shot inference, all models are developed using the training (70%) and validation (10%) set from each patient’s dataset. The final performance is assessed on the testing set (the remaining 20% held out dataset), and all reported metrics represent the mean values across all test samples. The comprehensive results for all 10 patients, measured by mean absolute and relative errors, are summarized in Table 4.1 and 4.2, along with the corresponding training time. The best-performing result for each patient in each metric is highlighted in bold.

As shown in Table 4.1 and 4.2, the fine-tuning approach demonstrates superior performance for most patients, such as S003–S007. To illustrate the advantage of each strategy, we analyze the results for a typical patient, say S003, as detailed in Table 4.1. First, zero-shot inference provides a strong baseline, achieving competitive performance without any additional training, which highlights the power of the pretrained foundation model for immediate deployment. Fine-tuning enhances performance across all metrics by training all model parameters on the new dataset, but at a cost of 31 seconds of training. In contrast, linear probing offers a highly efficient alternative. By updating only 0.36% of the total parameters (23,055 out of 6,337,039), it substantially saves the training time, but with a slight reduction in accuracy compared to the fine-tuning approach. The staged fine-tuning approach which combines these two methods, yields the strongest performance, although it also costs the longest training time. Finally, and most importantly, all strategies that take advantage of the pretrained model parameter values significantly outperform the training from a random initialization, underscoring the substantial benefit of transfer learning.

However, the specific result varies from patients to patients. We observe a notable contrast: while fine-tuning is the best-performing strategy for patient S001 (Table 4.1), the zero-shot inference achieves the highest accuracy for patients S008–S010 (Table 4.2). This underscores the complexity of transfer learning and may need further investigation into fine-tuning approaches. A potential explanation for this discrepancy in performance lies in the length of the time series, as we notice that patients S008–S010 have significantly shorter data records, each with approximately 1,000 data points This result seems to suggest the training outcome scales with the size of the training dataset in this particular project.

These findings suggest a practical guideline for applying transfer learning in a clinical setting. For instance, in the early stage of deploying the model to a new patient when data is scarce, leveraging the pretrained model via zero-shot inference appears to be the most plausible approach. Subsequently, as more data is collected over time, fine-tuning becomes a highly effective strategy for creating a personalized model. Compared to training a model from scratch, this approach yields higher accuracy and requires just a few extra seconds of training time. The choice between full and staged fine-tuning would depend on the desired trade-off between accuracy and computational cost. Overall, the longest training cost for retraining the model is less than 48s on a modestly configured laptop.

Next, we evaluate the model’s performance on a more challenging forecasting task, with the prediction horizon extended to 30 steps (30/30 setting). The results are presented in Tables 4.3 and 4.4. As shown, the training time is almost the same as the 30-15 setting, with a negligible increase of about one second. However, this extended horizon leads to an increase in the forecasting error, approximately twice that of the 30-15 setting in some cases. This finding is consistent with the conclusion reached in [4] about the foundation model, where a super linear increase in errors was observed for increasing the length of prediction horizons. Despite this increase, the worst-case relative error remains below 9.5% in one case, which is clinically acceptable.

**Table 4.3:**
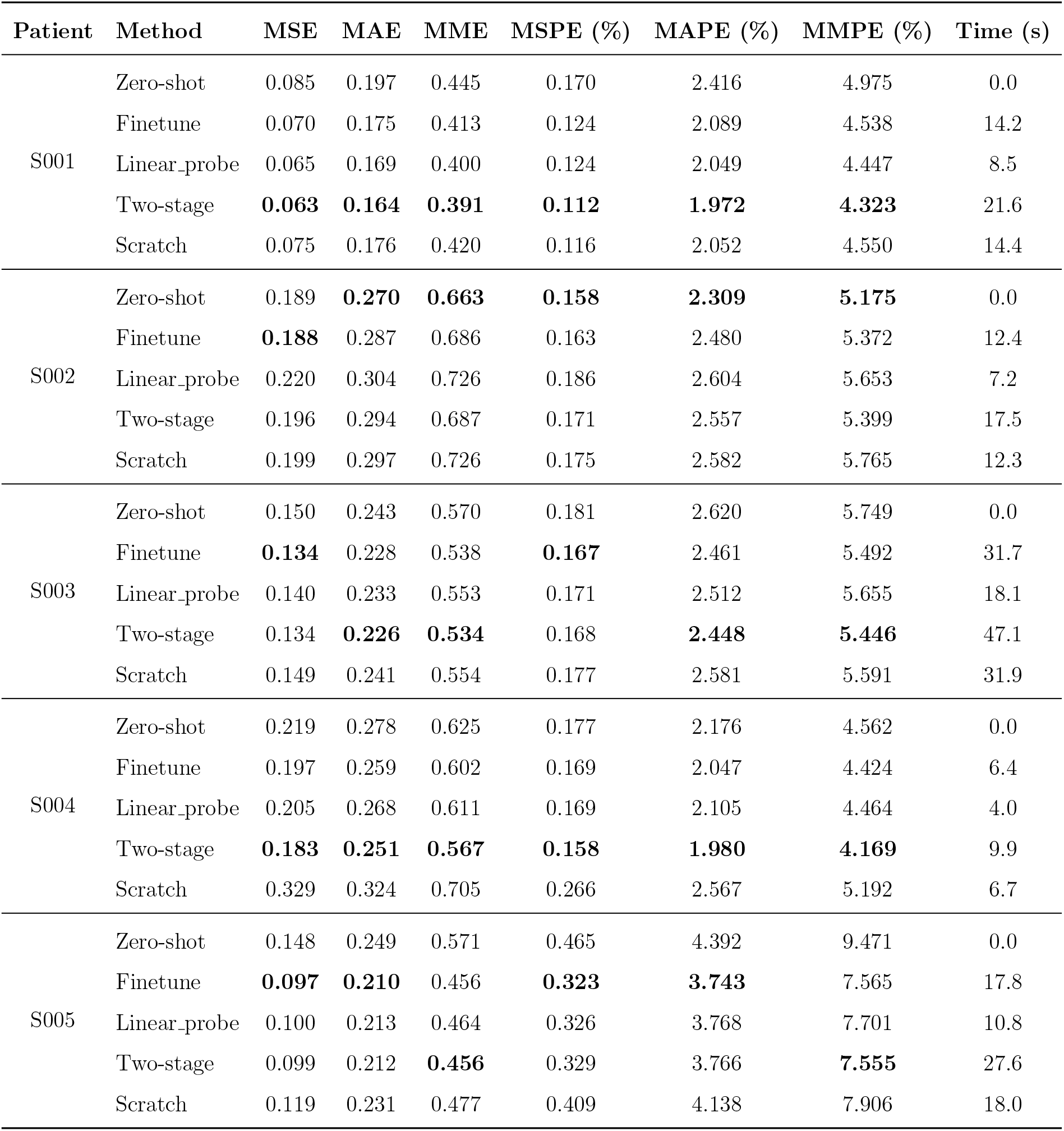
Forecasting performance and computational cost of five training and fine-tuning strategies for patients S001–S005. All experiments are conducted using a 30-step look-back window and a 30-step prediction horizon. The best result is highlighted in bold.

**Table 4.4:** Forecasting performance and computational cost of five training and fine-tuning strategies for patients S006–S010. All experiments were conducted using a 30-step look-back window and a 30-step prediction horizon. The best result is highlighted in bold.

| Patient | Method | MSE | MAE | MME | MSPE (%) | MAPE (%) | MMPE (%) | Time (s) |
| --- | --- | --- | --- | --- | --- | --- | --- | --- |
| S006 | Zero-shot | **0.135** | **0.237** | **0.558** | **0.095** | **1.911** | **4.288** | 0.0 |
|  | Finetune | 0.212 | 0.302 | 0.748 | 0.130 | 2.359 | 5.571 | 17.1 |
|  | Linear_probe | 0.240 | 0.323 | 0.786 | 0.146 | 2.514 | 5.852 | 10.1 |
|  | Two-stage | 0.221 | 0.315 | 0.767 | 0.135 | 2.452 | 5.705 | 26.4 |
|  | Scratch | 0.279 | 0.351 | 0.838 | 0.160 | 2.697 | 6.177 | 17.3 |
| S007 | Zero-shot | 0.112 | 0.226 | 0.510 | 0.052 | 1.545 | 3.434 | 0.0 |
|  | Finetune | 0.076 | 0.191 | 0.420 | 0.036 | 1.314 | 2.836 | 7.7 |
|  | Linear_probe | 0.088 | 0.204 | 0.457 | 0.041 | 1.403 | 3.081 | 5.1 |
|  | Two-stage | **0.073** | **0.187** | **0.411** | **0.034** | **1.285** | **2.776** | 12.0 |
|  | Scratch | 0.075 | 0.194 | 0.415 | 0.036 | 1.343 | 2.820 | 8.1 |
| S008 | Zero-shot | **0.065** | **0.168** | **0.354** | **0.040** | **1.318** | **2.753** | 0.0 |
|  | Finetune | 0.091 | 0.203 | 0.441 | 0.056 | 1.594 | 3.425 | 2.3 |
|  | Linear_probe | 0.081 | 0.191 | 0.402 | 0.050 | 1.501 | 3.122 | 1.7 |
|  | Two-stage | 0.100 | 0.213 | 0.471 | 0.061 | 1.669 | 3.658 | 3.5 |
|  | Scratch | 0.087 | 0.199 | 0.374 | 0.052 | 1.557 | 2.898 | 2.3 |
| S009 | Zero-shot | 0.200 | 0.292 | 0.662 | 0.146 | 2.474 | 5.446 | 0.0 |
|  | Finetune | 0.190 | **0.273** | **0.587** | 0.136 | **2.309** | **4.819** | 2.9 |
|  | Linear_probe | **0.182** | 0.277 | 0.616 | **0.134** | 2.355 | 5.063 | 1.9 |
|  | Two-stage | 0.200 | 0.277 | 0.592 | 0.144 | 2.346 | 4.855 | 4.5 |
|  | Scratch | 0.253 | 0.328 | 0.667 | 0.181 | 2.766 | 5.441 | 3.7 |
| S010 | Zero-shot | 0.081 | 0.194 | 0.435 | 0.043 | 1.407 | 3.045 | 0.0 |
|  | Finetune | 0.076 | 0.195 | 0.443 | 0.041 | 1.425 | 3.135 | 4.1 |
|  | Linear_probe | **0.069** | **0.182** | **0.417** | **0.036** | **1.322** | **2.932** | 2.9 |
|  | Two-stage | 0.099 | 0.213 | 0.478 | 0.056 | 1.574 | 3.433 | 6.4 |
|  | Scratch | 0.091 | 0.218 | 0.492 | 0.047 | 1.578 | 3.424 | 4.4 |

### 4.2 Digital twin and its workflow

To deploy the foundation model to a new patient to produce a digital twin, we devise the following workflow, which enables the digital twin to be not only built but also dynamically adapted for the new patient in a clinical setting quickly. For the digital twin, we must ensure the bidirectional information exchange between the digital twin and the patient and dynamical update whenever new patient data become available in real time. As illustrated in Fig. 4.1, the workflow comprises two steps: the initial deployment, bi-directional interaction, and dynamic update.

**Figure 4.1:**
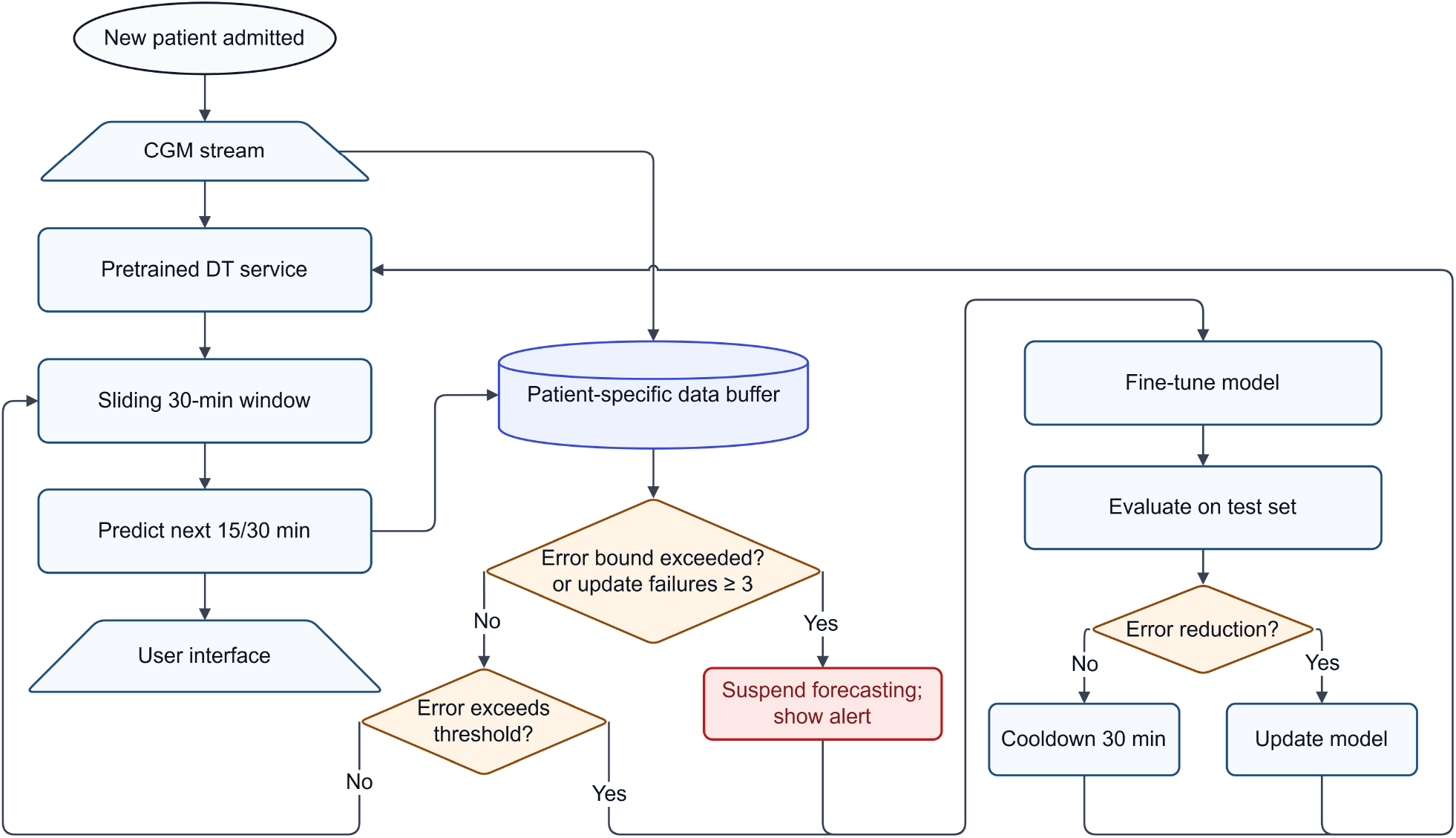
The workflow of a digital twin deployment and dynamic updating for a new patient.

- Initial Deployment (zero-shot inference) and bi-directional interaction: Upon the admission of a new patient, the pretrained foundation model is immediately deployed to perform the zero-shot inference. The model works on a rolling basis, continuously using the most recent 30 minutes of the patient’s glucose data to generate the next 15 or 30 minutes prediction. The predicted data are stored and benchmarked with the patient data retrospectively and in real time in the bi-directional interaction. This process provides users with immediate and constantly updated predictions, offering valuable initial clinical insight without any patient-specific training.
- Bi-directional Interaction and Dynamic Updating: This is a complex process consisting several loops and branches.
  - As time goes by, new ground-truth data is continuously collected from the new patient. For example, after 15 minutes, the model’s forecast can then be compared with the newly acquired measurements. This bi-directional interactive comparison generates a new labeled data sample (a prediction-truth pair), which is added to a patient-specific data buffer. This allows for real-time performance monitoring and creates a growing dataset for future adaptation.
  - A dynamic model update is triggered when the ensemble forecasting error exceeds a predefined threshold (e.g., ERE *≥ ϵ*, where *ϵ* is a user-specified threshold value). This initiates a rapid fine-tuning process on the new samples and simultaneously alerts the user about the model’s current performance. For safety, if the error surpasses a critical upper bound (e.g., ERE *≥ δ*, where *δ* is a user-defined error bound), the system automatically suspends its forecasting output, simultaneously alerts the user about the model’s current performance, and prioritizes model retraining. To ensure the performance and reliability of the updated model, the newly acquired data is partitioned into training, validation, and test sets, and the model is retrained using the fine-tuning method that users select.
  - Before re-deployment, the newly fine-tuned model is compared with the current model on this temporary test set. Only if the updated model outperformed the current model in predictive accuracy can it be deployed (e.g., MMPE), otherwise, the current model is retained to ensure system stability.
  - If the fine-tuned model fails this check, a cool-down period (e.g., 15 minutes) begins, during which the system pauses updating to collect more data. The system is also suspended after three consecutive failures, signaling the need for manual review and more comprehensive model retraining.
  - The entire updating process with any adverse disruptions is devised to be completed in less than one minute, ensuring the model continuously adapts to the patient’s evolving dynamics and provides clinicians with the most current, personalized forecasts.

Upon the arrival of new patient data, each sample is first normalized using the mean and variance derived from the source dataset. Following this initialization, the system employs a sliding window strategy: At any given time step *i*, the predictor generates a sequence of forecasts for the future *H* time steps. We denote this prediction as

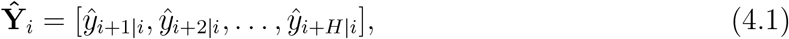

where ŷ_*t*|*i*_ represents the predicted value at target time *t* conditioned on given time point *i*. Due to the sequential nature of the sliding window, a specific target time point *t* is covered by multiple prediction windows generated at different past time points *t ™ k* (1 *≤ k ≤ H*). Consequently, for any target time *t*, we obtain a set of overlapping predictions, defined as

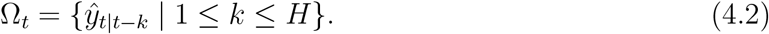

To ensemble these overlapping forecasts, we assign a time-decay weight *w*_*k*_ to each prediction based on its forecast horizon *k*,

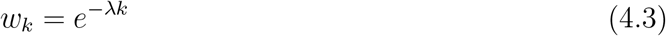

Here, *λ* (*λ ≥* 0) serves as a decay rate hyperparameter. A larger *λ* assigns significantly higher importance to recent forecasts, as we assume that predictions closer to the target time are generally more accurate. In this study, we set *λ* = 1. The final ensemble prediction value 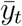 is obtained by taking weighted average of these forecasts:

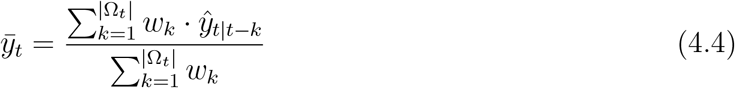

where |Ω_*t*_| denotes the number of predictions for time *t*, which equals *H* except during the initial warm-up phase. For real-time performance monitoring, we employ the Ensemble Relative Error (*ERE*) to compare with the ground truth. Let *y*_*t*_ denote the ground truth at time step *t*, the error is defined as

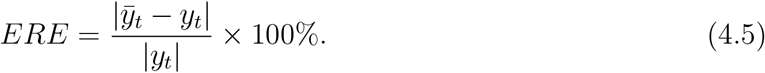

In our digital twin framework, *ERE* serves as the primary error metric for model adaptation. Specifically, a full fine-tuning update is triggered if this error exceeds *ϵ* = 5%, while forecasting is suspended if the error surpasses *δ* = 10%. The corresponding experimental results are detailed in Figures 4.2.

**Figure 4.2:**
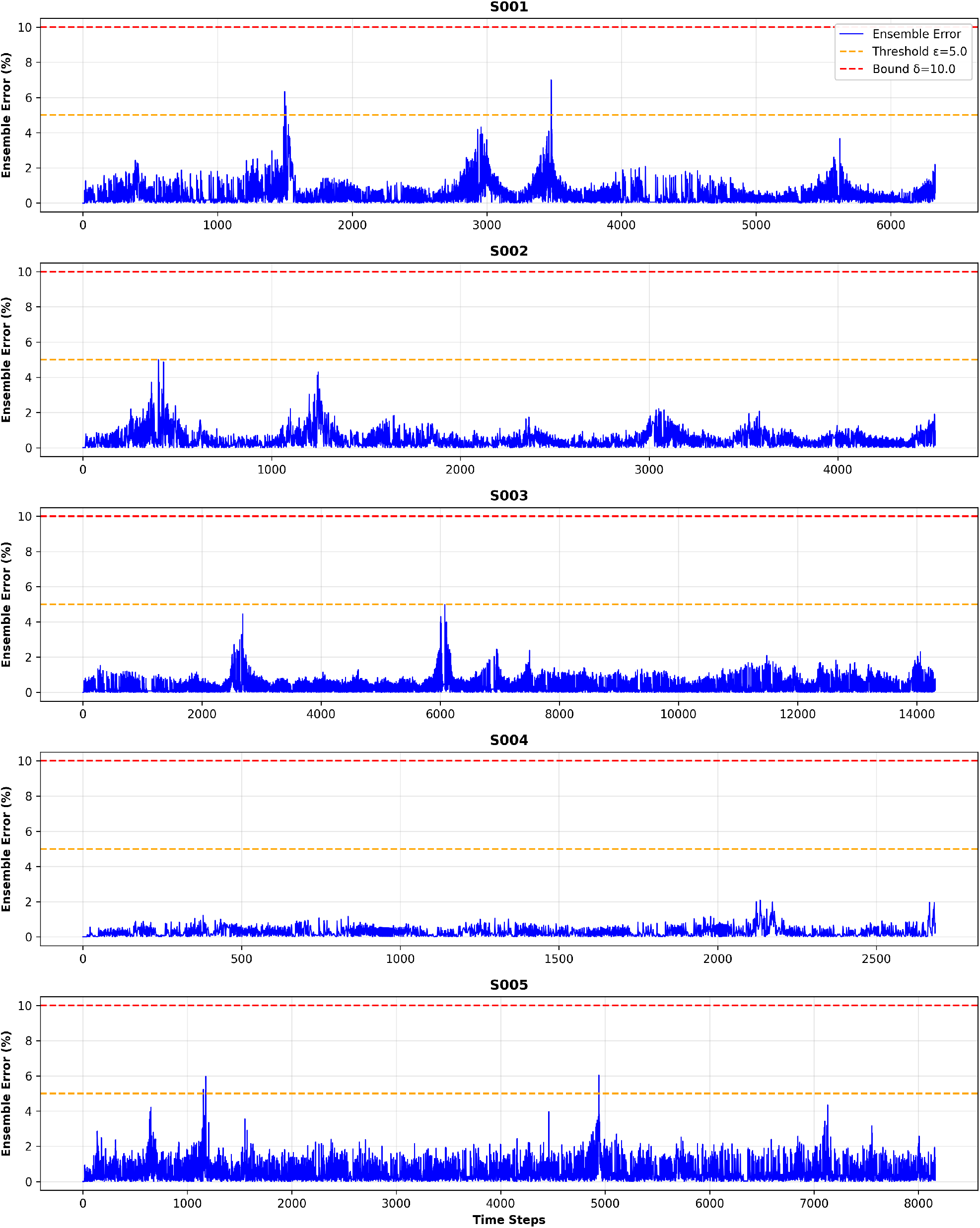
Dynamic error monitoring for five representative patients (S001–S005). The blue solid line represents the Ensemble Relative Error at each time step. The orange dashed line marks the model update threshold (*ϵ* = 5.0%), while the red dashed line indicates the safety suspension bound (*δ* = 10.0%).

As illustrated in the figure, the error for patient S001 remained below the threshold for most of the time. Model updates were triggered only twice, resulting in a rapid reduction in prediction error. In contrast, for patient S004, the error consistently stayed within the safety margins throughout the whole period, indicating that the pre-trained foundation model was sufficient for accurate forecasting without the need for fine-tuning.

The workflow has been applied to all 10 patients, where the digital twin has not shown any alert to stop forecasting during the time frame tested. The study result demonstrates that the failure of the digital twin workflow in the retrospective study of the 10 patient cases is an extremely rare event.

## 5 Discussion

The digital twin framework developed in this study demonstrates the potential of AI-enabled modeling for continuous glucose monitoring and forecasting in the intensive care unit (ICU). The ICU environment poses unique challenges—patients exhibit rapidly changing physiological states, high data variability, and multiple, interacting treatments. In such settings, real-time adaptive systems that integrate continuous data streams and update forecasts dynamically are critical. The proposed digital twin exemplifies such a system, providing a data-driven, continuously learning surrogate of the patient’s glucose metabolic state.

The integration of transfer learning within the digital twin’s workflow allows for rapid adaptation of a pretrained model to new patients with minimal amount of data. The results show that zero-shot inference delivers reliable initial forecasts immediately upon deployment, while fine-tuning strategies further refine the model as additional data accumulates. This adaptability is crucial in the ICU, where treatment decisions must often be made with sparse early data but can be optimized as patient-specific trends emerge. The ability to update models within seconds ensures clinical feasibility and supports continuous model personalization without interrupting care workflows.

Beyond glucose forecasting, this digital twin framework establishes a foundation for developing AI-assisted clinical decision support systems (CDSS) in critical care. By enabling predictive monitoring of key physiological variables, a digital twin can inform early intervention strategies, alert clinicians to impending hypo-or hyperglycemic events, and evaluate the potential outcomes of treatment adjustments before implementation. Such decision support complements, rather than replaces, clinician judgment—augmenting the decision-making process with quantitative, individualized predictions grounded in patient-specific data.

In the broader context of ICU management, digital twins can evolve into comprehensive physician aides and treatment planning tools. Coupled with multimodal data integration—such as hemodynamic, biochemical, and pharmacologic inputs—a multi-scale digital twin can simulate the effects of various therapeutic regimens, providing a virtual testbed for precision medicine. For instance, incorporating insulin dosing history, nutritional intake, and vasopressor use could allow the digital twin to project metabolic responses under alternative treatment scenarios. This capability aligns with the vision of “learning ICUs,” where AI systems continuously assimilate data, adapt to individual patients, and guide care optimization at both individual and population levels.

Importantly, the success of the presented digital twin framework underscores several design principles for digital twins in healthcare and medicine in general: (1) efficiency and deployability—the ability to initialize and fine-tune models on-site within seconds; (2) interpretability and trust—transparent representation of prediction confidence and model evolution; and (3) bidirectional communication—continuous feedback between digital twins and physical systems to enable self-correction and clinician-informed refinement.

Future work along the line will focus on extending this digital twin development paradigm to multi-functional digital twins that integrate heterogeneous data sources and support optimization of glycemic control and stability. Additionally, clinical validation with larger patient cohorts and diverse ICU populations will be essential for regulatory translation and integration into routine critical care practice.

Overall, the proposed digital twin demonstrates a promising step toward AI-empowered, personalized, and adaptive treatment and support systems in the ICU, transforming real-time data into actionable clinical insight.

### 5.1 Clinical Implications

The proposed digital twin framework provides a pathway toward real-time, personalized decision support in the ICU. By continuously integrating patient-specific data and adapting model predictions through transfer learning workflows, it can assist clinicians in anticipating critical glycemic events and tailoring treatment strategies dynamically. Its rapid deployability on readily available computing devices makes it suitable for bedside use, enabling seamless integration into existing ICU workflows without specialized infrastructure. Beyond glycemic control, the framework can be extended to other physiological systems—such as hemodynamics, respiratory function, or drug metabolism—offering a scalable foundation for multi-functional digital twins. Such digital twins can support early intervention, reduce treatment delays, and contribute to data-driven precision medicine. Ultimately, integrating digital twins into clinical decision pathways has the potential to enhance patient safety, optimize therapeutic efficacy, and improve outcomes in complex, high-acuity care environments.

## 6 Conclusion

This study presents a digital twin framework for continuous glucose monitoring and forecasting in septic patients admitted to the ICU, demonstrating the feasibility and clinical value of AI-enabled modeling in critical care. By leveraging transfer learning workflows, the proposed digital twin effectively adapts a pretrained/foundation glucose forecasting model to new patients with minimal data and computational cost. The results show that the model can be deployed within seconds, providing accurate real-time glucose predictions and supporting dynamic model updating as patient data evolves.

The findings highlight that zero-shot inference offers reliable early-stage forecasts when patient data are limited, while fine-tuning strategies yield improved personalized predictions as more data are collected. This progressive adaptation makes the digital twin an efficient and practical tool for continuous monitoring and individualized treatment planning. The ability to integrate real-time data streams and generate predictive insights positions the system as a valuable AI-aided decision support tool, capable of alerting clinicians to potential glycemic instabilities and evaluating therapeutic strategies before implementation.

More broadly, this work illustrates how digital twins can serve as in-silico companions to critically ill patients—bridging continuous sensing, predictive modeling, and clinical decision-making. Future extensions will focus on developing multimodal digital twins that integrate additional physiological, imaging, clinical record, and biochemical variables to provide a more holistic representation of patient dynamics. As such systems evolve, they hold the potential to transform the ICU into a continuously learning environment—where personalized, data-driven care plans are refined in real time to improve patient outcomes and optimize resource use.

In summary, the proposed digital twin framework establishes a scalable foundation for AI-driven precision medicine in critical care, demonstrating how real-time analytics and adaptive modeling can be integrated into clinical workflows to support timely, informed, and individualized treatment decisions.

## Acknowledgments

The authors would like to acknowledge the contribution made by Dr. Hua Jiang’s group to this study by sharing with us their dataset collected. Xiang Cao, Jianguo Hou, Xinhang Wei, and Qi Wang’s research is partially supported by NSF awards DMS-2038080 and OIA-2242812; Chao Cai and Qi Wang’s research is partially supported by an SC GAIN-CRP award. The funders played no role in study design, data collection, analysis and interpretation of data, or the writing of this manuscript.

## Contributions

X.C. and Q. W. conceived the study. X.C. conceptualized the model and conducted the analyses. J.H. and X.W. performed the literature review to inform model parameters. X.C., J.H., and X.W. validated the model structure and assumptions and provided feedback on parameter selection. X.C. and Q.W. interpreted the results and prepared the first draft. Q.W. and C.C. revised the manuscript. Q.W. and C.C. secured funding for the project. All authors contributed to the interpretation of the findings, reviewed the manuscript, and approved the final manuscript.

## Data availability

The datasets used and/or analyzed during the current study are available from the corresponding author on reasonable request.

## Code availability

The underlying code for this study and training/validation datasets is not publicly available but may be made available to researchers on reasonable requests from the corresponding authors.

